# Epidemiology and Predictors of Fluoroquinolone Resistance in ESBL-Producing *Escherichia coli*: Implications for Empirical Therapy in Mexico

**DOI:** 10.64898/2026.04.21.26351439

**Authors:** Abigail Gallardo Mejia, Jorge Almeida

## Abstract

Urinary tract infections (UTIs) are among the most common infectious diseases worldwide, with *Escherichia coli* being the predominant uropathogen. The increasing prevalence of extended-spectrum beta-lactamase (ESBL)-producing strains and their association with fluoroquinolone resistance pose a significant challenge to empirical therapy, particularly in community settings.

The aim of this study was to determine the epidemiology and predictive factors associated with ESBL-producing *E. coli* and its concomitant fluoroquinolone resistance in community-acquired clinical isolates.

A retrospective cross-sectional study was conducted analyzing 244 clinical *E. coli* isolates. Demographic and microbiological data were collected, including age, sex, sample type, and antibiotic susceptibility. Associations between variables and ESBL production were assessed using Pearson’s chi-squared test, and odds ratios (ORs) with 95% confidence intervals (CIs) were calculated.

Of the isolates, 165 (68%) were ESBL-producing. A significant association was observed between age group and ESBL production (p < 0.001), with the highest frequency in the 20–39 age group. Most ESBL-positive isolates were obtained from women (73%), although odds ratio (OR) analysis suggested a non-significant trend toward a higher probability in men (OR = 1.29; 95% CI: 0.72–2.31). High rates of fluoroquinolone resistance were identified among the ESBL-producing isolates, with 30% resistance to levofloxacin and 35% to ciprofloxacin (p < 0.001). Urine samples showed the highest concentration of ESBL-positive isolates, with a significant association between sample type and resistance (p < 0.001).

The high prevalence of ESBL-producing *E. coli* and its concomitant resistance to fluoroquinolones highlight a critical challenge for the empirical treatment of urinary tract infections in Mexico, underscoring the need to strengthen antimicrobial use management and local surveillance strategies.

## Introduction

Urinary tract infections (UTIs) are one of the most common pathological conditions in both community and hospital settings(1), affecting over 150 million individuals each year. These infections are associated with considerable morbidity and encompass a broad spectrum of clinical manifestations(2). Urinary tract infection (UTI) poses a serious health issue for all age groups from neonates to geriatric age groups. Although men and women can become infected, majority of the women experience urinary tract infections once in their lifetime(3,4). The etiology of UTI is also affected by underlying host factors that complicate UTI, such as age, diabetes, spinal cord injury, or catheterization(5). UTIs are the second most common type of nosocomial infection. UTIs can be divided into three categories: acute pyelonephritis, acute cystitis, and asymptomatic bacteriuria (6).

Urinary tract infections are caused by a range of pathogens, but most commonly by *Escherichia coli, Klebsiella pneumoniae, Proteus mirabilis, Enterococcus faecalis and Staphylococcus saprophyticus, Pseudomonas aeruginosa, Staphylococcus aureus* and Candida spp. The microbial etiology of urinary infections has been regarded as well established and reasonably consistent. *Escherichia coli* remains the predominant uropathogen (UPEC) (80%) isolated in acute community-acquired uncomplicated infections, followed by Staphylococcus saprophyticus (10% to 15%) (7,8).

UPEC is a highly adaptive pathogen which presents significant treatment challenges rooted in a complex interplay of molecular factors that allow UPEC to evade host defences, persist within the urinary tract, and resist antibiotic therapy(9). The effectiveness of conventional treatments against *E. coli* has been seriously compromised by the global emergence of Extended-Spectrum Beta-Lactamase (ESBL)-producing strains (10). The acquisition and expression of (ESBL), mainly CTX-M, TEM and SHV, is the main β-lactam resistance in these strains. Moreover, mutations in the genes encoding DNA gyrase (gyrA/gyrB) and topoisomerase IV (parC/parE), as well as the acquisition of genes such as qnrA/B, may lead to fluoroquinolone resistance (11).

In 2023, the Ministry of Health of Mexico reported that UTIs are the second leading cause of morbidity, with a total of 3,365,799 identified cases(12). In this global landscape of resistance, local epidemiological surveillance is essential to guide empirical therapy and mitigate the spread of resistant strains. At the regional level, there is a lack of up-to-date information that integrates temporal dynamics and demographic factors within the community, as well as the presence of enzymes such as ESBLs. The objective of this study was to determine the epidemiology and predictive factors of extended-spectrum beta-lactamase (ESBL)-producing *Escherichia coli* exhibiting multidrug resistance in a series of clinical isolates collected over one year. The prevalence of co-resistance to fluoroquinolones-presence of ESBL was studied and the association between age groups, sex and quarterly distribution was assessed, to provide evidence to support the need for stricter management protocols in the face of the high rate of resistance observed in community-origin isolates.

## Materials and Methods

### Study Design and Context

A retrospective, observational, cross-sectional study was conducted to analyze clinical isolates of *Escherichia coli* obtained over one year (2024). The study focused on community-acquired isolates to assess the prevalence of extended-spectrum beta-lactamase (ESBL) production and cross-resistance to fluoroquinolones.

### Data Collection and Bacterial Isolates

A total of 244 antibiogram reports from community-origin clinical samples with cultures positive for *E. coli* were collected. For each isolated strain, demographic and microbiological data were extracted, including patient age, sex, sample type, quarter of isolation, and susceptibility profiles to various antibiotics.

Microbiological studies for species determination, antibiotic sensitivity patterns, and phenotypic resistance pattern markers were performed using the Beckman Coulter Microscan WalkAway Plus automated method. The data from patients were encrypted to protect personal information. Antimicrobial susceptibility testing (AST) was performed using amikacin (AMK), amoxicillin– clavulanic acid (AMC), ampicillin (AMP), ampicillin–sulbactam (SAM), cefoxitin (FOX), ceftazidime (CAZ), ceftriaxone (CRO), ciprofloxacin (CIP), ertapenem (ETP), gentamicin (GEN), imipenem (IPM), levofloxacin (LVX), meropenem (MEM), tetracycline (TCY), tigecycline (TGC), piperacillin– tazobactam (TZP), trimethoprim–sulfamethoxazole (SXT).

### Statistical Analysis

Ages were segmented into 10-year intervals, forming the following age groups: 0–9, 10–19, 20–29, 30–39, 40–49, 50–59, 60–69, 70–79, and 80–89 years. For the Pearson’s chi-square test, the ages were regrouped, and the groups were segmented as follows: 0–19, 20–39, 40–59, and ≥60, in order to meet the statistical assumptions of the test. The detailed segmentation by decade was used for descriptive analysis. For inferential analysis; the age groups were regrouped to ensure the validity of the statistical test. A descriptive analysis was performed, calculating absolute and relative frequencies (n and %) for the categorical variables. For the age variable, measures of central tendency (mean and mode) and measures of dispersion (standard deviation and range) were calculated.

To evaluate the association between age groups and the presence of ESBLs, Pearson’s chi-square test was applied, considering a p-value < 0.05 as statistically significant.

## Results

Data on total crops, age, gender and crop type can be found in Table 1.

**Table 1.** Demographic characteristics.

| CHARACTERISTICS OF THE PATIENTS | n=244 | % |
| --- | --- | --- |
| **Gender** |  |  |
| Male | 65 | 27 |
| Female | 179 | 73 |
| **Groups** |  |  |
| CCV | 57 | 23 |
| FAR | 7 | 3 |
| GEN | 28 | 11 |
| URO | 152 | 62 |
| **BLEE** |  |  |
| POSITIVE | 165 | 68 |
| NEGATIVE | 79 | 32 |
| **MDR** |  |  |
| POSITIVE | 195 | 80 |
| NEGATIVE | 49 | 20 |
| **AGE** |  |  |
| 0-9 | 3 | 1 |
| 10-19 | 16 | 7 |
| 20-29 | 42 | 17 |
| 30-39 | 44 | 18 |
| 40-49 | 47 | 19 |
| 50-59 | 38 | 16 |
| 60-69 | 27 | 11 |
| 70-79 | 2 | 1 |
| 80-89 | 25 | 10 |

### -Age distribution

The mean age of the study population was 44.7 years, with a mode of 35 years, the 20–39 age group being the modal range.

The highest frequency of isolates was observed in the 30–39 age group (44 cases) and 40–49 age group (47 cases), followed by the 20–29 age group (42 cases). The extreme age groups showed the lowest frequency.

### -Antibiotic resistance and presence of ESBL

Of the 244 clinical isolates analyzed, 165 (68%) were ESBL producers.

For penicillin-derived beta-lactams, 69% of isolates (n=168) showed resistance to ampicillin, 38% (n=93) to ampicillin/sulbactam, 35% (n=85) to amoxicillin/clavulanic acid, and 15% (n=36) to piperacillin/tazobactam. For cephalosporins, 70% (n=170) were resistant to cefotaxime, 68% (n=165) to ceftriaxone, 64% (n=155) to ceftazidime, and 32% (n=79) to cefepime. For carbapenems, there was resistance of 25% (n=60) to imipenem, 26% (n=63) to ertapenem, and 21% (n=51) to meropenem. Among fluoroquinolones, the isolates showed resistance of 27% (n=66) to levofloxacin and 30% (n=72) to ciprofloxacin. For aminoglycosides, resistance was 35% (n=86) to gentamicin and 41% (n=100) to amikacin. Resistance to tetracyclines was 47% (n=115) for tetracycline and 38% (n=93) for tigecycline; finally, for trimethoprim/sulfamethoxazole, resistance was 45% (n=111) table 2, Figure 1.

**Table 2.** Percentages of antibiotic resistance out of all *E. coli* isolates.

| ANTIBIOTICS | RESISTANT | SENSITIVE |
| --- | --- | --- |
| Ampicillin | 69% | 31% |
| Ampicillin/Sulbactam | 38% | 62% |
| Amoxicillin/Clavulanic Acid | 35% | 65% |
| Ceftazidime | 64% | 36% |
| Ceftriaxone | 68% | 32% |
| Cefepime | 32% | 68% |
| Piperacillin/Tazobactam | 15% | 85% |
| Meropenem | 21% | 79% |
| Ertapenem | 26% | 74% |
| Imipenem | 25% | 75% |
| Levofloxacin | 30% | 70% |
| Ciprofloxacin | 35% | 65% |
| Gentamicin | 35% | 65% |
| Amikacin | 41% | 59% |
| Tetracycline | 47% | 53% |
| Tigecycline | 38% | 62% |
| Trimethoprim/Sulfamethoxazole | 45% | 55% |

**Figure 1.**
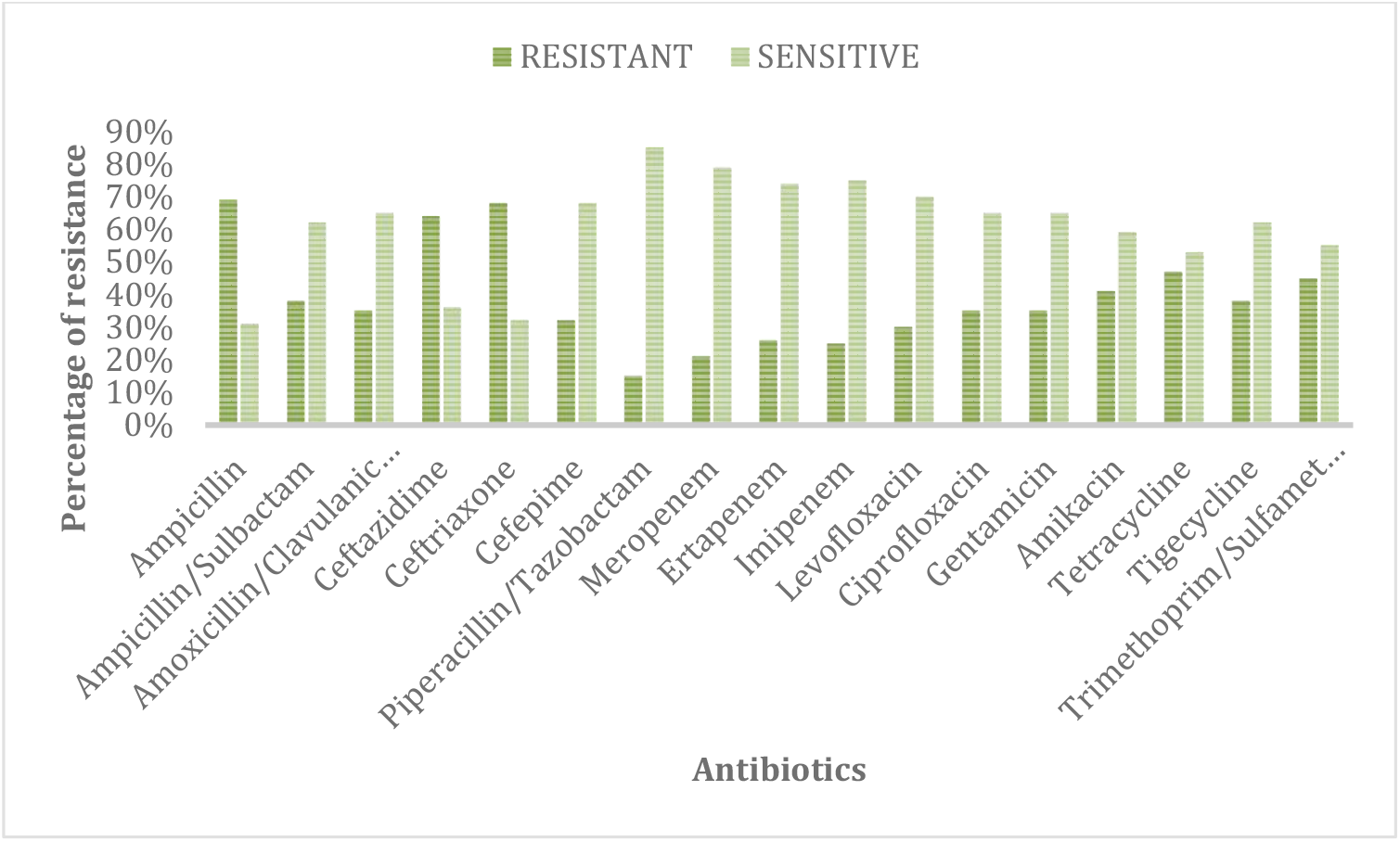
Percentages of antibiotic resistance out of all *E. coli* isolates.

### -Association between age group and ESBL production

When age was grouped into clinical intervals (0–19, 20–39, 40–59, and ≥60 years), the 20–39 age group had the highest number of ESBL-positive isolates (64 cases), followed by the 40–59 age group (64 cases) with the same number of cases.

Pearson’s chi-square test showed a statistically significant association between age group and ESBL production (χ^2^, p = 1.9 × 10^-5^; p < 0.001), indicating that the distribution of ESBL production was not independent of age.

### -Distribution by sex

Of the total positive ESBL isolates, n=120 (73%) were from women and n=45 (27%) from men. The average age was higher in women (73.36 years) compared to men (26.64 years). The association between ESBL production and sex was evaluated using Pearson’s chi-square test, finding a statistically significant association (χ^2^ = 3.45277E-34; p < 0.001). The analysis using odds ratio showed a trend towards a higher probability of ESBL production in men (OR = 1.29; 95% CI: 0.72– 2.31); however, the confidence interval included unity.

### -Fluoroquinolone resistance in ESBL-producing isolates

Of the total ESBL-positive isolates, n=50 (30%) showed resistance to levofloxacin, while n=57 (35%) showed resistance to ciprofloxacin. Overall, a high frequency of fluoroquinolone resistance was observed among the ESBL-producing isolates. The association between ESBL production and resistance to levofloxacin and ciprofloxacin was evaluated using Pearson’s chi-square test, revealing a statistically significant association (χ^2^ = 3.73363E-29; p < 0.001).

### -Association between sample type and ESBL

When grouping the types of clinical isolates (CCV, FAR/GEN, URO), it was observed that the urine culture sample type (URO) had the highest number of ESBL-positive isolates (104 cases). The Pearson chi-square test showed a statistically significant association between sample type and the presence of ESBL-positive isolates (χ^2^, p = 2.97 × 10^-^78; p < 0.001).

The results show a high prevalence of ESBLs, particularly in young and middle-aged adults, with a statistically significant association between age and ESBL production, as well as high resistance to fluoroquinolones, which represents a significant challenge for therapeutic management.

### -Temporal distribution

The highest frequency of MDR isolates was observed during the first quarter (55 cases), followed by the second quarter (50 cases). Regarding ESBLs, the highest values were recorded during the third and fourth quarters (38 cases each).

## Discussion

In the present study, *Escherichia coli* was confirmed as the main uropathogen, according to some international reports that indicate that this microorganism represents more than 60% of isolations in urinary tract infections (UTIs), both in the community and hospital settings (13).

Of the 244 clinical isolates analyzed, n=165 (68%) were extended-spectrum β-lactamase (ESBL) producers, representing a high prevalence. Epidemiological studies report regional variability in the prevalence of these strains. In Latin American countries, prevalences of around 40.1% have been reported in Peru, while in Mexico, the figures can reach up to 50%, depending on the population studied and the isolation method used (14). The high proportion examined in this study suggests a significant endemic problem of antimicrobial resistance, typical of contexts where antibiotic use is frequent or not adequately regulated through rational use programs.

Regarding the demographic characteristics of the target population, the mean age was 44.7 years, with a modal group between 10 and 19 years, although a high frequency of isolates was also observed in the 30–39 and 40–49 age groups. This demonstrates that the presence of ESBL-producing strains is not limited solely to elderly populations. However, several epidemiological studies have shown that the probability of resistant isolates tends to increase proportionally with age and with the presence of comorbidities or previous antibiotic exposures (15).

For statistical analysis, the age groups were restructured into four categories (0–19, 20–39, 40–59, and ≥60 years), allowing the application of the chi-square test and examination of the association between age and ESBL production. The results showed that the 20–39 age group had 64 ESBL-positive isolates, with a statistically significant association (p < 0.001) observed between age and the presence of this resistance mechanism. Similar findings have been reported in clinical studies that identify age and a history of antimicrobial exposure as factors associated with the isolation of ESBL strains (16).

Regarding sex, 71% of ESBL-positive isolates were from women and 29% from men, which is consistent with the classic epidemiology of urinary tract infections. Women have a greater predisposition to developing UTIs due to anatomical and physiological factors, which increases the frequency of urinary *E. coli* isolates in this population group (17).

Regarding the antimicrobial resistance profile, high resistance to fluoroquinolones was observed among the ESBL-positive isolates. Specifically, 30% of the strains showed resistance to levofloxacin and 35% to ciprofloxacin. These results are comparable to epidemiological surveillance studies that have reported resistance rates exceeding 70% in ESBL-producing strains, confirming the close association between these two resistance mechanisms (18).

Research conducted in Brazil has demonstrated that ESBL-producing *E. coli* strains can exhibit fluoroquinolone resistance rates between 82.1% and 89.6%, significantly higher values compared to non-ESBL-producing strains (18). Similarly, European studies have documented that fluoroquinolone resistance in uropathogens with an ESBL phenotype often exceeds 60%, which considerably hinders the use of these antibiotics as initial empirical therapy (19).

Recent reports in Mexico show that the prevalence of ESBL in community-acquired UTIs ranges from 21-56%, and co-resistance to fluoroquinolones in ESBL+ strains reaches up to 80-90% in some hospital and community studies (20).

Additionally, when grouping the types of clinical isolates analyzed (CCV, FAR/GEN, and URO), it was observed that the urine culture sample (URO) had the highest number of ESBL-positive isolates, with 152 cases. This finding is consistent with the high frequency of E. coli as the main etiological agent of urinary tract infections.

To evaluate the relationship between the type of clinical sample and the presence of ESBLs, Pearson’s chi-square test was applied, which showed a statistically significant association between both variables (χ^2^; p = 2.97 × 10^-78^; p < 0.001). These results indicate that the distribution of ESBL-producing strains is not random among the different sample types, but rather shows a higher concentration in urinary isolates.

International literature shows that, although there are differences between regions, antimicrobial resistance in uropathogens shows an increasing trend globally, positioning itself as one of the main threats to public health (21).

The coexistence of ESBL production and fluoroquinolone resistance has been associated with a higher risk of therapeutic failure, increased hospital stays and higher health costs, especially when initial empirical treatment is inadequate (22).

In this context, the Mexican Clinical Practice Guideline No. 39.0 Urinary Tract Infection, Unspecified Site, Diagnosis and Treatment of Acute, Uncomplicated Urinary Tract Infection in Women (ISBN: 978-607-8270-14-9) explicitly states the first-line treatment as trimethoprim-sulfamethoxazole, nitrofurantoin as a second-line choice, ampicillin and amoxicillin with clavulanic acid, as well as the use of fluoroquinolones, mainly ciprofloxacin, for complications and empirical treatment while awaiting urine culture results. This information is quoted verbatim below; “In non-pregnant women of “any age” with signs and symptoms of acute lower urinary tract infection, treatment should be with TMP/SMZ (160/800mg; twice a day for 3 days) as first choice or with Nitrofurantoin (100 mg twice a day for 7 days) as second choice. There is a high rate of ampicillin resistance in microorganisms that cause pyelonephritis, and a high rate of recurrence is observed in women treated empirically with beta-lactamases. The exception is when the causative agent is Gram-positive; in these cases, amoxicillin and amoxicillin with clavulanic acid can be used. Treatment should be considered for 14 days orally, parenterally, or both. A clinically evident response should be present 48 to 72 hours after starting the antibiotic. A follow-up urine culture should be taken 14 days after completing treatment.

Gram-positive clusters (likely *staphylococcus*) can be initially treated with cephalosporins. In other cases, beta-lactamases are not recommended. The first-line drug is quinolones for 14 days. In areas with low resistance to TMP/SMZ, it is an acceptable alternative. In all cases, whether the patient is hospitalized or not, 14 days of treatment must be completed.

In patients without severe symptoms or comorbidities and who can tolerate oral intake, the patient should be informed of the risks and benefits of outpatient treatment compared to inpatient treatment. If their physical condition permits and the patient agrees, outpatient treatment will be given with follow-up at 48 and 72 hours; a culture should be taken, and treatment with ciprofloxacin 500 mg every 12 hours should be initiated for 14 days. A follow-up culture should be taken 2 weeks after completing the treatment. Monitor progress at 48 to 72 hours. Second choice TMP SMX (160/800 mg) every 12 hours for 14 days”(23).

However, the data in the present study show a high percentage of resistance to both trimethoprim-sulfamethoxazole and recommended beta-lactams. Additionally, although 30 to 35 percent of strains are resistant to fluoroquinolones, there is a high number of strains with ESBL production, which directly causes therapeutic failure against cirprofloxacin through induction mechanisms.

In the case of fluoroquinolones, resistance in E. coli frequently occurs through mutations in the gyrA gene and, less frequently, in the gyrB gene, which catalyzes ATP-dependent DNA supercoiling. Other mechanisms of *E. coli* resistance to quinolones and fluoroquinolones include efflux pumps and reduced drug accumulation in the bacteria due to changes in the purine protein (24). Numerous studies have revealed that mutations in a small region at the N-terminal end of gyrA (amino acids 67 (Ala-67) to 106 (Gln-106)) confer resistance to quinolones and fluoroquinolones, known as the quinolone resistance-determining region (QRDR).

The production of β-lactamase enzymes is the most common mechanism of bacterial resistance. Extended-spectrum β-lactamase (ESBL)-producing bacteria often exhibit multidrug resistance because, in most cases, the genes associated with other resistance mechanisms are also located on the same plasmid that contains the ESBL-encoding genes (25).

The same clinical practice guideline, in its updated 2024 version, suggests not using fluoroquinolones, especially ciprofloxacin, as first-line treatment, unlike its 2009 edition. This change is not due to resistance patterns, but rather to the adverse effects and side effects this class of drugs causes in patients. On the other hand, trimethoprim-sulfamethoxazole continues to be suggested as a first-line strategy, as the information and evidence cited in this clinical practice guideline is based on studies published in the Argentine population in 2016 (26), disregarding all studies in the Mexican population, particularly relevant to this study and its geographic area, the Toluca Valley. This omission has led to therapeutic failure with the drugs suggested as first-line treatments.

On the other hand, a high percentage of carbapenemases was detected, ranging from 21% to 26% in meropenem and ertapenem, respectively. This finding is consistent with a multicenter study conducted in Mexico, which demonstrated an increase in carbapenemase-producing strains of *E. coli* exhibiting bla NDM, as well as a high prevalence of AmpC-type beta-lactamases(27).

## Conclusion

The present study demonstrates a high prevalence of ESBL-producing Escherichia coli (68%) in clinical isolates of community origin in the Toluca Valley, Mexico. This figure significantly exceeds reports from previous years, confirming an alarming trend toward multidrug resistance (MDR) in the extra-hospital setting.The statistical analysis identified age as a critical predictive factor for the presence of ESBL-producing strains (p < 0.001), with a high incidence in both the 0–19 and 40–59 age groups. Furthermore, the predominance of these infections in the female population (71%) aligns with the classic epidemiology of UTIs but highlights the increasing complexity of treating common infections in women of all ages.A major finding of this research is the high rate of co-resistance to fluoroquinolones among ESBL-positive isolates, reaching 30% for levofloxacin and 35% for ciprofloxacin. These levels of resistance render fluoroquinolones—and many first-line beta-lactams—ineffective as empirical therapy.Finally, our results reveal a significant discrepancy between local microbiological reality and the recommendations of the Mexican Clinical Practice Guidelines (GPC). The reliance on outdated or non-regional evidence in national protocols leads to an increased risk of therapeutic failure. Therefore, it is imperative to update clinical guidelines based on local epidemiological surveillance to ensure the rational use of antibiotics and the success of treatment protocols in Mexico.

## Data Availability

The data is available upon request

## Acknowledgments

Victoria, for her invaluable motivation

